# Unraveling the Link Between Cognition and Motor Impairment and Activity After Stroke: A Longitudinal Study

**DOI:** 10.64898/2026.04.29.26352027

**Authors:** Reut Binyamin Netser, Adi Lorber-Haddad, Noy Goldhamer, Hadar Idan, Adi Tayer Yeshurun, Gil Meir, Keren Pollack, Tamar Mizrahi, Simona Bar Haim, Yogev Koren, Lior Shmuelof

**Affiliations:** Department of Industrial Engineering and Management, Ben-Gurion University of The Negev, Be’er-Sheva, Israel; Adi Negev - Nahalat Eran Rehabilitation Center, Ofakim, Israel; Department of Occupational Therapy, Ben-Gurion University of the Negev, Beer Sheva, Israel; The School of Brain Sciences and Cognition, Ben-Gurion University of The Negev, Be’er-Sheva; The Lillian and David E. Feldman Research Center for Rehabilitation Sciences, Adi Negev - Nahalat Eran Medical Center; Department of Physical Therapy, Ben-Gurion University of the Negev, Beer Sheva, Israel

**Keywords:** stroke recovery, cognitive impairment, motor recovery, neurorehabilitation, longitudinal study, functional outcomes

## Abstract

**Background:** Stroke leads to both motor and cognitive impairments that can substantially limit daily activities and independence. Although these impairments are often treated separately in rehabilitation, growing evidence suggests they are interconnected. Understanding how cognitive and motor impairments relate to one another is essential for developing more effective, integrated rehabilitation strategies.

**Objective:** This longitudinal study addressed three key questions: (1) Do motor and cognitive impairments co-occur after stroke? (2) Does cognitive ability influence motor recovery? (3) Are cognitive and motor recovery trajectories associated?

**Methods:** We followed 148 individuals in the subacute phase of stroke, assessing them at 1 and 3 months post-stroke. Cognitive function was measured using the Montreal Cognitive Assessment (MoCA) and the clock drawing test. Motor impairment was assessed using the Fugl-Meyer Assessment (FMA) and grip strength. Activity was evaluated using the Action Research Arm Test (ARAT), 10-Meter Walk Test (10MW), and Timed Up and Go (TUG).

**Results:** At one month post-stroke, cognitive and motor impairment and activity levels were not correlated, although strong within-domain correlations were observed. Baseline cognitive ability did not predict motor impairment recovery. However, improvements in cognitive ability from 1 to 3 months were moderately correlated with gains in motor activity measures (r = 0.22–0.29, p < 0.05).

**Conclusions:** Although cognitive and motor impairments may arise independently after stroke, their recovery processes appear partially linked. These findings underscore the importance of addressing both domains in rehabilitation and advancing understanding of shared mechanisms that support recovery across functional systems.

## Introduction

Stroke frequently results in motor impairments that significantly limit activity, functional independence, and quality of life. The most common motor impairment is hemiparesis, which can affect both gross and fine motor control (Langhorne et al., 2009). Recovery of motor impairments is typically incomplete: only 12% of people with stroke (PwS) achieve full restoration of the upper extremity abilities within six months post-stroke (Nijland et al., 2010). Nearly half of the PwS continue to experience residual motor impairments three months after the event (Hatem et al., 2016). Persistent motor impairment is associated with abnormal activity, reduced participation, decreased independence, and long-term disability (Kim, 2022).

In addition to motor impairments, stroke frequently leads to cognitive impairments. Cognitive impairments occur in up to 72% of PwS (Sexton et al., 2019). They may involve a wide range of sub-domains, including attention, memory, executive functions, planning, and problem-solving (Nys et al., 2007). The prevalence and severity of cognitive impairments vary depending on lesion location, size, and pre-stroke cognitive status (Béjot et al., 2020; Zhao et al., 2017), with persistent impairments observed in a substantial proportion of PwS, months or even years after the event (Munthe-Kaas et al., 2020). Cognitive impairments have been linked to reduced functional independence, poorer quality of life, and lower likelihood of successful return to community living (Heruti et al., 2002; Mok et al., 2004; Ones et al., 2009). Traditionally, impairments in the motor and cognitive domains have been addressed separately in rehabilitation settings. However, a growing corpus of evidence suggests that motor and cognitive impairments are interconnected (Binyamin-Netser et al., 2023; Lingo VanGilder et al., 2020).

What is the nature of the association between cognitive and motor impairments after stroke? The reported association could result from damage to shared underlying neural substrates of cognitive and motor control. For example, stroke following MCA occlusion is known to be associated with both weakness, aphasia, and disorientation (Gao et al., 2020). A recent study showed that 28% of PwS show impairment in cognition and motor function after stroke, which is related to lesion volume and age (Einstad et al., 2022). The interconnection between domains could be an outcome of their overlapping neural representations, resulting in overlapping manifestations of impairments or from the contribution of cognitive abilities to motor performance in clinical measures of motor activity and participation (Chen et al., 2013). Our first hypothesis is that motor impairments and motor activity limitations will appear together with cognitive impairments.

Another form of association is causal. Cognitive impairment may affect PwS’s daily activities and their level of engagement in rehabilitation, thereby indirectly affecting their motor recovery. Cognitive processes, such as executive functions and memory, are known to be impaired after stroke (Nys et al., 2007). Indeed, a longitudinal study showed that impairment of executive functions was associated with poorer motor recovery (Mancuso et al., 2023). Our second hypothesis is that cognitive impairments affect motor recovery.

Lastly, the recovery in the two domains may be correlated due to shared recovery mechanisms. General aspects that affect the ability to recover may lead to a correlation between impairment and activity limitation recovery in cognitive and motor domains. For example, age was found to be an influential factor in both motor and cognitive recovery after stroke (Yoo et al., 2020). The third hypothesis is that motor and cognitive recovery are associated.

This study aims to identify indications of the suggested associations between stroke effects on the cognitive and motor domains in a longitudinal study. We focus on associations between primary impairment assessments in each domain, as a measure of mechanistic overlap, and on the association of cognitive impairments and activity limitations as a possible measure of functional overlap. Understanding the association between domains is crucial for the development of effective neurotechnological interventions that typically rely on high cognitive abilities.

## Methods

### Subjects

148 PwS in the sub-acute phase were recruited to this longitudinal study. The study was conducted at the Adi Negev - Nahalat Eran rehabilitation center (Israel) in collaboration with The Lillian and David E. Feldman Research Center for Rehabilitation Sciences and was approved by the Regional Ethical Review Board at Adi Negev, Israel (Approval Number ADINEGEV-2023_106). All patients with stroke (PwS) provided a written informed consent before enrollment. Inclusion criteria were: age ≥18 years, ischemic or hemorrhagic stroke (hemispheric or brainstem) confirmed by CT or MRI, first-ever stroke or a previous stroke with no UE weakness before the second incident, more than 1 week and less than 6 weeks after the stroke, and intact ability to understand and sign a consent form. Exclusion criteria: history of physical or neurological condition that interferes with study procedures or assessment of motor function (e.g., severe arthritis, severe neuropathy, Parkinson’s disease). Dependence in daily activities before the stroke with documented motor impairments.

Data produced in the present study is available upon reasonable request to the authors.

#### Procedure

Participants were assessed on two occasions: at baseline (approximately 1 month after stroke) and three months following stroke onset (±2 weeks). During each session, participants were evaluated for cognitive and motor deficits.

### Tools

UE and LE motor impairment evaluation was divided into assessments of impairment (the underlying problem with body function) and activity (the ability to perform functional tasks). UE motor impairment was assessed using the Fugl-Meyer Assessment (FMA) of the upper-limb (UE FMA), a widely used clinical measure that assesses motor impairment, including movement, coordination, and reflexes. Higher scores indicate better function (maximum score: 66) (Duncan et al., 1983; Fugl-Meyer et al., 1975), and grip strength (using microFET dynamometer). Grip strength scores were standardized to account for age and sex differences and normalized (z-scores) (Biasini et al., 2023; Bohannon, 2006). UE motor activity was assessed using the Action Research Arm Test (ARAT), which is used to measure the ability to perform functional tasks with the arm and hand, such as grasping, gripping, pinching, and gross movement (maximum score: 57) (Yozbatiran et al., 2008). LE motor impairment was assessed using the lower extremity sub-section of the FMA (LE FMA) (maximum score: 34). LE motor activity was assessed using the 10 Meter Walk (10MW), which measures walking duration over a 10-meter distance. Durations were converted to speed (10/duration) and is reported in m/sec (Collen et al., 1990; Severinsen et al., 2011; Wolf et al., 1999). This conversion enabled us to include participants unable to walk with a meaningful score (0 m/sec). Another tool we used was Time Up and Go (TUG), which is a common measure for functional mobility. (Flansbjer et al., 2005). While duration is the common outcome of this test, we used speed (6/duration) following the same logic above. Cognitive impairment was assessed using the Montreal Cognitive Assessment (MoCA) as a primary measure (Chiti & Pantoni, 2014; Nasreddine et al., 2005) and the clock drawing test as a secondary measure (CDT) (Pinto & Peters, 2009; Rouleau et al., 1992; Shulman, 2000). These cognitive screening instruments were chosen given their clinical applicability, and since we did not have specific hypotheses regarding cognitive sub-domains. In several cases (15%), due to a change in the protocol at an early stage, the Mini-Mental State Examination (MMSE) (Cumming et al., 2013; Folstein et al., 1975) was used instead of the MoCA. SWEET 16 cognitive evaluation (Fong et al., 2011) was used for subjects with no education (0 years of study). The scores of MMSE and SWEET 16 were transformed to MoCA scores using established matrices (Fong et al., 2011; Helmi et al., 2016).

### Data analysis

The number of PwS differed across examinations and time points due to several reasons, such as fatigue, lack of motivation to continue the study, and difficulties associated with returning to the second evaluation (Table 1). Correlation analyses were computed only for PwS with both examinations. Comparison of impairment levels (t-tests) was computed for the entire group of PwS.

**Table 1:**
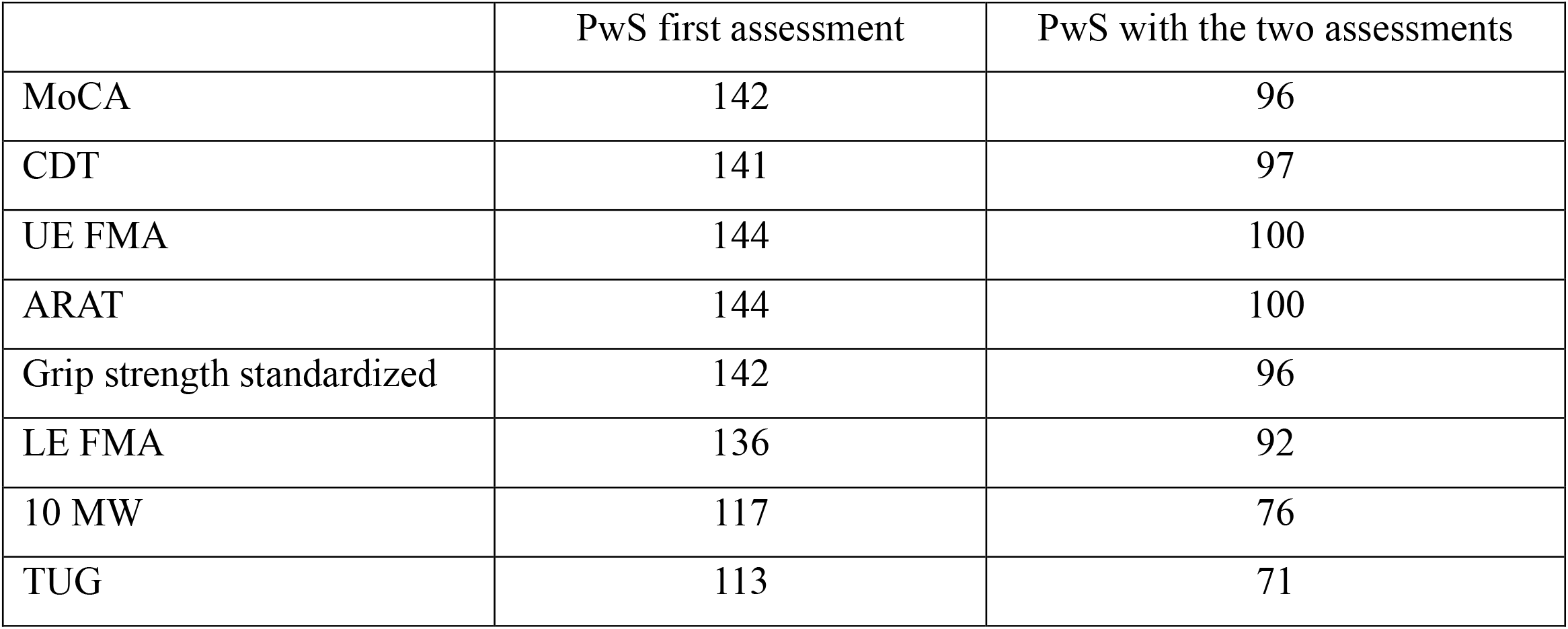
The number of examinations for each assessment in baseline and with two assessments.

|  | PwS first assessment | PwS with the two assessments |
| --- | --- | --- |
| MoCA | 142 | 96 |
| CDT | 141 | 97 |
| UE FMA | 144 | 100 |
| ARAT | 144 | 100 |
| Grip strength standardized | 142 | 96 |
| LE FMA | 136 | 92 |
| 10 MW | 117 | 76 |
| TUG | 113 | 71 |

Comparisons within and across domains were separated into correlation of MoCA with motor impairment and activity limitation assessments and recovery measures. For each domain, a primary measure of impairment and activity limitation was examined. Motor impairment in both UE and LE domains was assessed using the FMA as a primary measure. A secondary UE impairment measure was grip force. For activity limitation measures, ARAT was the primary (and only) measure of UE, and 10mw was the primary measure of LE. Correlations of secondary measures were conducted only if the primary measure of that domain showed a significant effect.

### Statistical analysis

All statistical calculations were performed using the Statistical Package SPSS 29 (IBM SPSS Statistics).

#### Evaluation of impairment in each outcome measure

To test whether the patients were impaired at the baseline evaluation (∼1 month after stroke), we used one sample t-test of each outcome measure compared to the maximal score an established threshold values (MoCA – 26; CDT – 10; UE FMA – 66; ARAT – 57; Grip strength – Z score; LE FMA – 34; 10MW – 1.2 m/s and TUG – 0.43 m/s).

#### Recovery analyses

To test the effect of time (first Vs. Second evaluation), we used the Wilcoxon Signed Rank Test for each of the outcome measures.

#### Impairment correlation analyses

To test the association between impairment scores across domains, values of each outcome in the baseline evaluation were correlated using Spearman’s correlation test (MoCA, CDT, UE FMA, ARAT, Grip strength, LE FMA, 10MW, and TUG).

#### Association between cognitive impairment and motor recovery

To test whether the level of cognitive impairment affect motor recovery, we tested the correlation between the baseline cognitive impairment (MoCA and clock drawing) and the change in motor impairment level (the score of the first assessment minus the score in the second assessment) in all of our motor outcomes (UE FMA, ARAT, Grip strength, LE FMA, 10MW and TUG), using Spearman’s correlation test.

#### Association between recovery rates in the different domains

To test whether recovery in the different domains was associated, we used Spearman’s correlation test between the change in examination scores (the scores in the second assessment minus the scores in the first assessment) in the different domains. Correlation analyses were made between all the tested parameters (MoCA, clock drawing, UE FMA, ARAT, Grip strength, LE FMA, 10MW, and TUG).

## Results

148 PwS were recruited to this study (mean age: 67.51 years, SD 9.85; 38 females). Many failed to complete all assessments at both time points (Table 1). The mean duration from stroke to the baseline assessment was 23.27 days (SD 10.45), and the mean duration from stroke to the second assessment was 91.94 days (SD 9.12). Upon enrollment, the study population showed impairments in cognitive, upper extremity, and lower extremity assessments, as indicated by submaximal scores (Table 2). Recovery, as indicated by improved assessments in the second timepoint (3 months) was observed in all assessments (Figure 1, Table 2).

**Table 2:** Mean score in each tested parameter in the first time point (1 month), the second time point (3 months), and the median delta between the two time points (3 months-1 month). In the parentheses, the Standard deviation score, the Z value of the Wilcoxon signed-rank test, and the p-value.

|  | Mean scores at 1 month (SD, p<sup>a</sup>) | Mean scores at 3 months (SD) | Median delta between timepoints (Z, p<sup>b</sup>) |
| --- | --- | --- | --- |
| MoCA | 18.96 (5.44, <.001) | 21.57 (5.80) | 2.00 (4.99, <.001) |
| CDT | 7.25 (2.93, <.001) | 8.21 (2.58) | 0.00 (3.27, <.001) |
| UE FMA | 45.33 (22.15, <.001) | 49.98 (19.17) | 4.00 (6.16, <.001) |
| ARAT | 35.64 (22.95, <.001) | 40.95 (20.83) | 2.00 (6.39, <.001) |
| Grip strength standardized | -2.64 (1.63, <.001) | -2.59 (1.54) | 0.26 (4.41, <.001) |
| LE FMA | 22.65 (8.66, <.001) | 25.23 (7.00) | 2.00 (4.47, <.001) |
| 10 MW | 0.67 (0.33, <.001) | 0.80 (0.36) | 0.22 (6.31, <.001) |
| TUG | 0.34 (0.19, <.001) | 0.41 (0.21) | 0.13 (6.75, <.001) |

<sup>a</sup> One sample t test indicating if the patients were impaired in this measure.
<sup>b</sup> Wilcoxon signed-rank test between the two timepoints, indicating if there is recovery.

**Figure 1:**
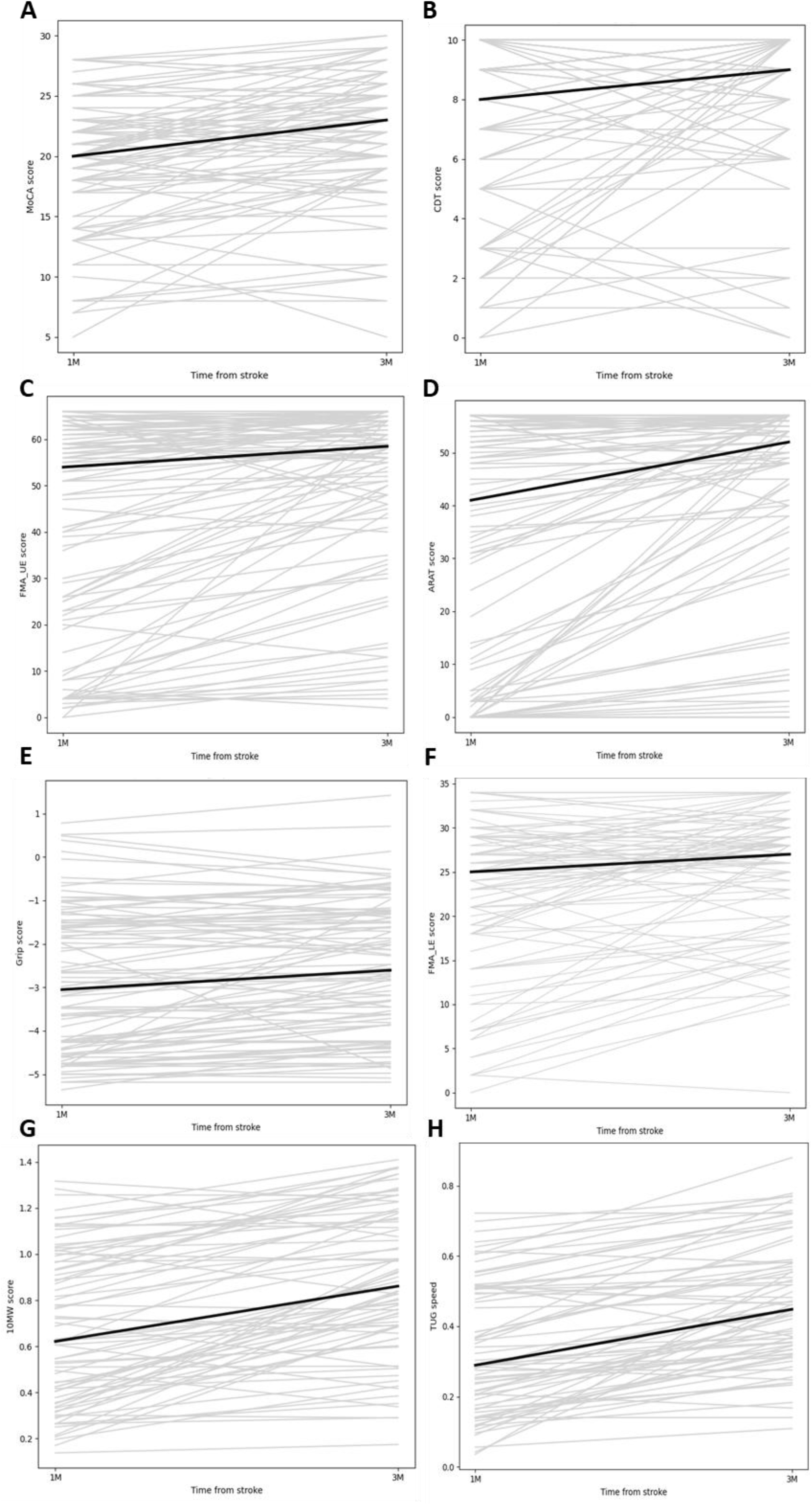
Recovery plots between the first (1 month) and second assessment (3 months). All gray lines refer to patients. Black line is the median of recovery across patients. A. MoCA B. CDT C. UE FMA D. ARAT E. Grip (standardized) F. LE FMA G. 10 MW H. TUG.

First, we tested the association between cognitive and motor impairments. Results revealed no correlation between the primary measures of each domain (Figure 2). There was a correlation between upper and lower extremity motor impairment (FMA UE and LE, r_s(131)_= 0.59, p<0.001) and activity limitation (ARAT and 10mw, r_s(112)_= 0.41, p<0.001). Importantly, within each domain, a strong correlation was observed between impairment assessments (FMA UE and grip, r_s(138)_= 0.80, p<0.001), activity limitation assessments (10MW and TUG r_s(103)_= 0.45, p<0.001) and between impairment and activity limitation assessments (FMA UE and ARAT, r_s(141)_= 0.86, p<0.001; FMA LE and 10MW, r_s(107)_= 0.43, p<0.001) (red frames in Figure 2).

**Figure 2:**
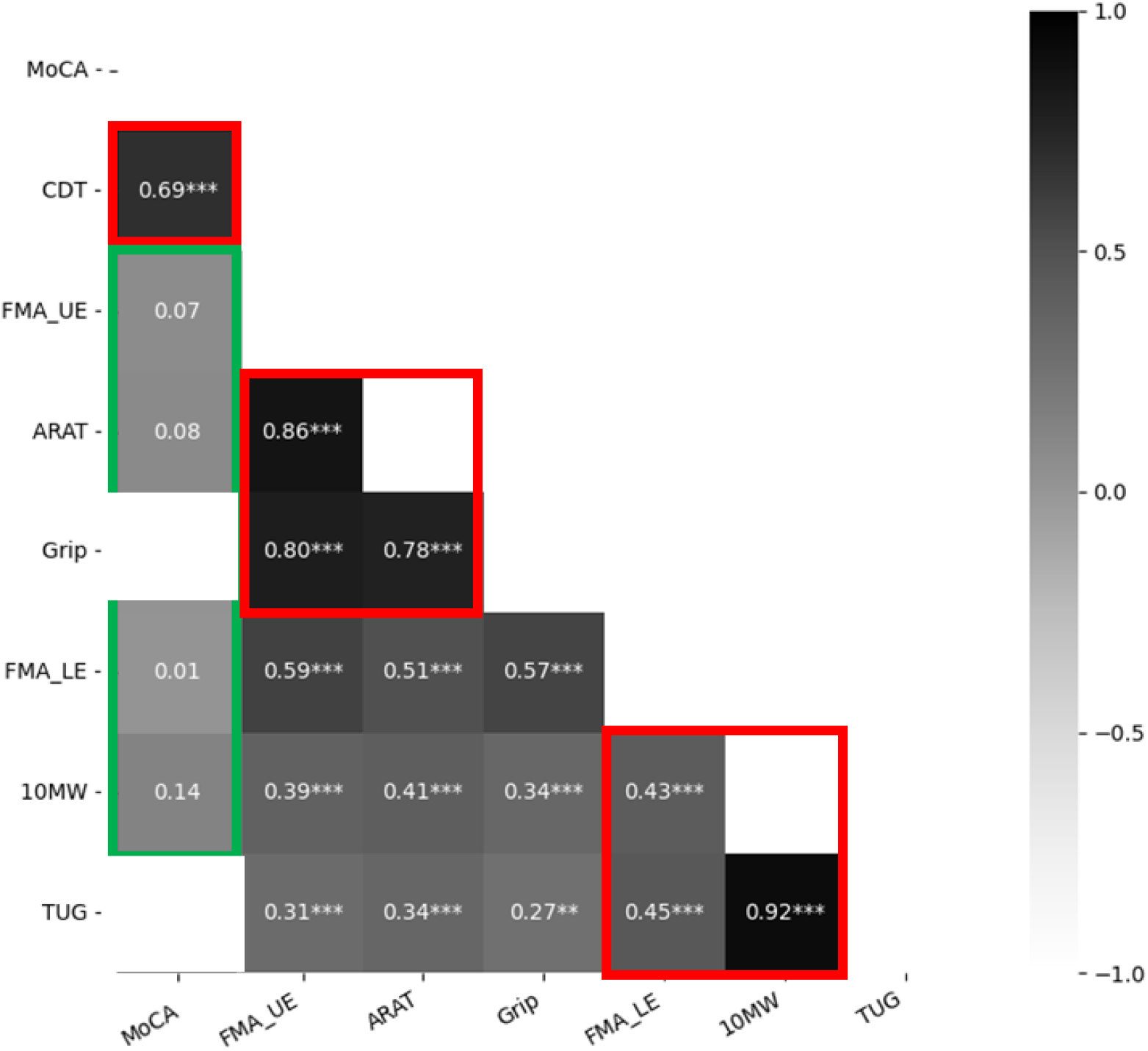
Heatmap of Spearman’s correlation coefficients among cognitive, upper extremity (UE), and lower extremity (LE) motor measures collected one-month post-stroke. Each cell contains the correlation coefficient (r), while the cell color represents the strength of the correlation-darker colors indicate higher r (hence stronger relationships), and brighter tones indicate lower r (hence weaker relationships). Asterisks represent significance, *p<0.05, **p<0.01, ***p<0.001. Red frames indicate correlations within domains, and green frames indicate correlations between domains (cognitive measures vs. UE and LE motor measures).

The second hypothesis was that cognitive function after stroke may affect motor recovery. The correlations between cognitive impairment at baseline and primary measures of impairment and activity limitation motor recovery (UE and LE) were not significant (Table 3).

**Table 3:** Spearman correlation coefficient between cognitive level (score in 1 month) and motor recovery (delta between 1 and three months). The p value is presented in parenthesis.

| | ΔUE FMA | ΔARAT | ΔLE FMA | Δ10MW |
| --- | --- | --- | --- | --- |
| MoCA (1M) | .06 (.53) | .01 (.92) | .08 (.46) | .02 (.90) |

Last, we examined the hypothesis that the recovery processes between the domains are associated. Cognitive recovery (ΔMoCA) was not correlated with recovery of motor impairment in the UE and LE. Cognitive recovery was correlated with activity limitation recovery of the UE (ΔARAT, r_s(93)_=0.22, p=0.04) and of the LE (Δ10MW, r_s(70)_=0.26, p=0.03; ΔTUG, r_s(65)_=0.29, p=0.02), suggesting a link between cognitive and functional motor gains (Figure 3). All domains showed internal correlations (Table 4). Motor domains also showed intercorrelations in recovery of impairment (ΔUE and ΔLE FMA r_s(89)_=0.35, p<0.001; ΔGrip and ΔLE FMA, r_s(86)_=0.25, p=0.02) but not in activity limitation recovery (ΔARAT and Δ10MW, r_s(73)_=0.16, p=0.18).

**Table 4:** Spearman correlation coefficient between cognitive and motor recovery (delta between 1 and three months). In the parenthesis, the p value.

| | ΔMoCA | ΔCDT | ΔFMA_UE | ΔARAT | ΔGrip | ΔFMA_LE | Δ10MW | ΔTUG |
| --- | --- | --- | --- | --- | --- | --- | --- | --- |
| ΔMoCA | — | .49 (<.001) | .06 (.57) | .22 (.04) | .18 (.10) | .10 (.37) | .26 (.03) | .29 (.02) |
| ΔCDT | | — | | .16 (.121) | | | .24 (.04) | .15 (.24) |
| ΔFMA_UE | | | — | .66 (<.001) | .56 (<.001) | .35 (<.001) | .17 (.15) | |
| ΔARAT | | | | — | .52 (<.001) | .12 (.27) | .16 (.18) | |
| ΔGrip | | | | | — | | .23 (.05) | .32 (.01) |
| ΔFMA_LE | | | | | | — | .34 (.004) | .40 (<.001) |
| Δ10MW | | | | | | | — | .71 (<.001) |
| ΔTUG | | | | | | | | — |

**Figure 3:**
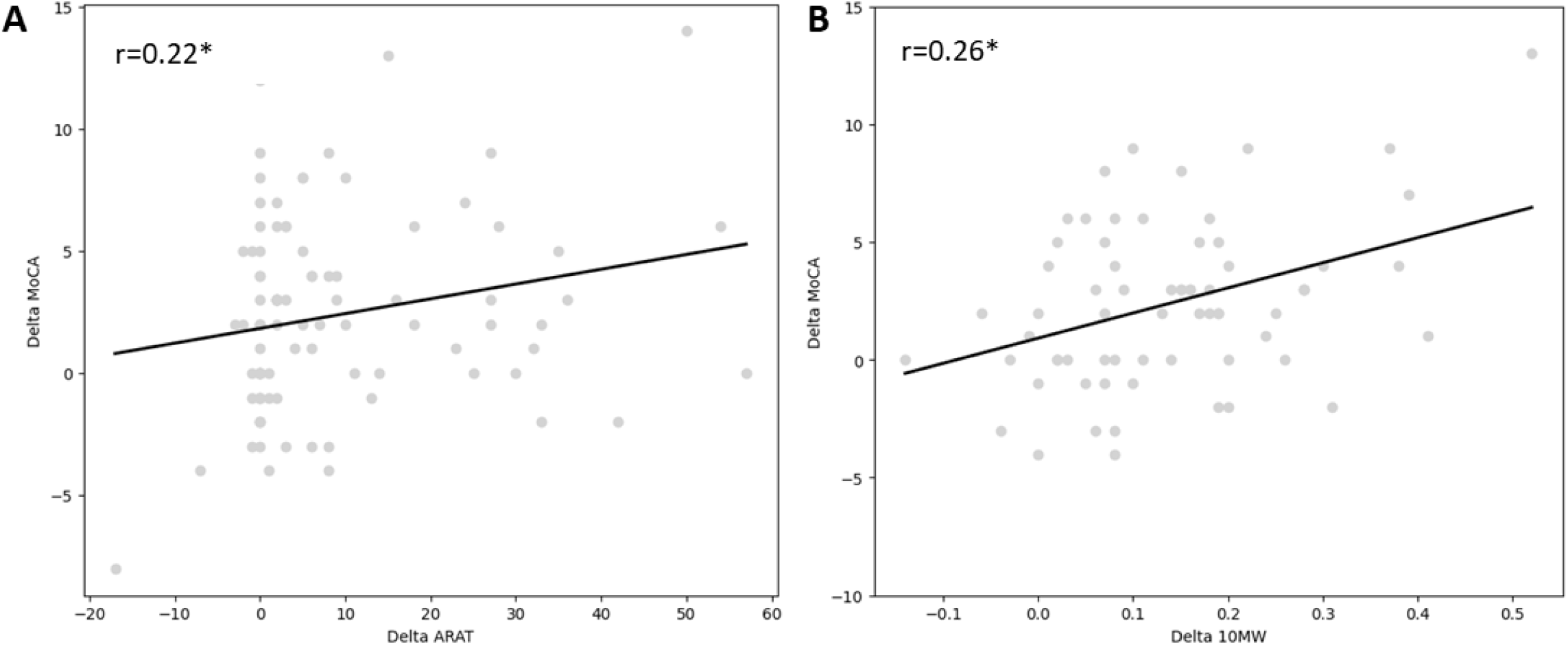
Spearman’s correlation among change scores (Δ3-1) for cognitive, upper extremity (UE), and lower extremity (LE) motor activity limitation measures assessed between one and three months post-stroke. Each dot refers to a patient, and the black line is a fit line. A. delta MoCA vs. delta ARAT. B. delta MoCA vs. delta 10 MW.

## Discussion

This longitudinal study examined the relationships between cognitive and motor impairments following stroke and recovery in the subacute stage. Our results show that motor and cognitive impairment estimates were not correlated one month after stroke, and that cognitive impairment estimates are not predictive of motor recovery. Our results point to an association between cognitive and motor activity limitation recovery.

Contrary to our initial hypothesis, we did not observe an association between cognitive and motor impairment estimates, despite the presence of within-domain correlations. These findings differ from those of Einstad et al. (2021) that reported an association between MoCA scores and grip force in a sample of 463 chronic PwS. This discrepancy may stem from the difference in sample size, in primary outcome measures (grip vs FMA UE in our study) or from possible long term interactive effects of cognition and activity in the chronic stage. Our results suggest that while stroke leads to cognitive and motor impairments, these deficits do not necessarily manifest together, likely due to their separate neural representations.

Our second hypothesis, that baseline cognitive function would influence motor recovery, was also not supported. Despite expectations and previous longitudinal reports linking executive dysfunction with poorer motor outcomes (Mancuso et al., 2023), our data did not reveal significant associations between early cognitive ability and subsequent upper or lower extremity recovery. Several factors may explain this discrepancy. First, much of the early post-stroke recovery is driven by spontaneous recovery (Cramer, 2008; Stinear & Byblow, 2014; Zeiler & Krakauer, 2013), which are largely time-dependent and less influenced by behavioral engagement. Second, structured inpatient rehabilitation programs may mitigate cognitive deficits through external scaffolding and guided practice, thereby buffering against the potential negative impact of impaired cognition on motor outcomes (Cicerone et al., 2011; Lanctôt et al., 2020). Third, the cognitive estimates we used are screening tests that may be insensitive to the cognitive impairments that directly affect motor rehabilitation, such as inhibition control (Demeyere et al., 2015). Together, these mechanisms may have masked a causal relationship between cognition and motor recovery in our cohort.

Interestingly, our third hypothesis - that recovery profiles across domains would be correlated - was partially supported. We found medium associations between improvements in cognitive performance and gains in functional motor measures.

These results suggest that although cognitive and motor impairments may arise independently, their recovery processes may be driven by the same mechanism, potentially reflecting shared biological processes such as neuroplasticity mediated by growth factors like BDNF (Ploughman et al., 2009) or external influences, including family support, environmental enrichment, and patient motivation (Fang et al., 2017; Khan et al., 2016).

### Limitations

Several limitations should be noted. First, our sample consisted of inpatients receiving intensive multidisciplinary care, which may not generalize to community-dwelling populations or outpatient rehabilitation settings. Second, our cognitive estimates (MoCA and CDT) are screening tools and do not capture specific cognitive subdomains or pathologies such as neglect and apraxia, which may show stronger links with motor impairments and recovery (Buxbaum et al., 2004). Third, our population is likely to consist of a subpopulation that may show dependencies between cognitive and motor functions. Our findings, therefore, reflect the general population of PwS that admitted to our facility with as few exclusions as possible. More elaborate selection criteria, such as studying subjects with cognitive impairments, may reveal dependencies between cognitive and motor functions. Alternatively, stratification of subjects based on the level of initial impairment or the location of brain damage may reveal subgroups with cognitive–motor dependencies (Lee & Kim, 2025).

## Conclusion

Our findings suggest that while cognitive and motor impairments may be manifested independently in the early subacute stage after stroke, their recovery is partially correlated. These results underscore the importance of identifying and enhancing shared recovery mechanisms and contextual influences. A deeper understanding of the dependencies between cognitive and motor sub-domains may allow the development of individualized cognitive-motor rehabilitation protocols.

## Data Availability

Data produced in the present study is available upon reasonable request to the authors.

## Acknowledgments

This research was supported by the Israeli Science Foundation (Grant 1244/22 to LS) and the Lillian and David E. Feldman Research fund.

